# Prevalence and Prescribers’ Knowledge of Psychotropic Polypharmacy in the Bono, Bono East, and Ahafo Regions, Ghana

**DOI:** 10.1101/2023.08.17.23294246

**Authors:** James Dumba, Antwi Joseph Barimah, Mohammed Mohammed Ibrahim, Solomon Saka Allotey, Semefa Alorvi, William Appertey, Luke Sopaal, Frank Acheampong, Benjamin Pulle Niriwa

**Author notes:** **Corresponding Author:** Antwi Joseph Barimah.

## Abstract

The use of psychotropic medications for treating simple and complex psychological and emotional problems is common and relevant among prescribers. This, therefore, come with the tendency to prescribe many medications to a patient on a single visit due to varied reasons. The study, therefore, sought to ascertain the prevalence and prescribers’ knowledge of psychotropic polypharmacy. A quantitative, descriptive study was conducted using a simple random approach to select and review the prescription records of three hundred and ten (309) patients’ folders retrospectively within seven (7) mental health units across the study area to ascertain the prevalence of Psychotropic Polypharmacy (PP). Fifty-eight (58) prescribers were also selected using probability method (simple random) to respond to the study questionnaire. Psychotropic Polypharmacy was prevalent (66.0%) in the study area with antipsychotic polypharmacy (74.0%) being the common form with the co-prescription of Chlorpromazine (CPZ) + Hadol (70.0%) being frequent. This was more predominant among male patients (78.0%) than female patients (22.0%). Prescribers had high (82.8%) knowledge of Psychotropic Polypharmacy and the majority (68.9%) disagreed that the practice of Psychotropic Polypharmacy should be promoted. The continuous training of prescribers (i.e. mental health officers) on rational prescriptions would help reduce the prevalence of Psychotropic Polypharmacy.

## Introduction

Psychotropic medications have been used over the period worldwide as a means of relieving or curing the signs and symptoms of mental illnesses/disorders, maintaining stability, prevention of relapses, and complications. Studies conducted have shown increasing degrees of psychotropic drug use and the concurrent use of multiple psychotropic medicines (polypharmacy) among children overall between 1–5years as well as in children with Autism Spectrum Disorders (ASD).^1^

The World Health Organization (WHO) has highlighted polypharmacy among prescribers as one of the major areas of focus in its Third Global Patient Safety Challenge, Medication without Harm.^2^ Polypharmacy is known to occur when the possible advantages of multiple medicines outweigh the detrimental consequences of a patient’s sheer number of medications prescribed. The incidence of polypharmacy is not simply about the number of drugs a patient uses, but also about the efficacy, usefulness, and possible harm of the drug either individually or in combination.^3^ Polypharmacy is a worldwide concern in both physical and mental health facilities.^4^ Globally, there appears to be a recounted high predominance of psychiatric or psychotropic polypharmacy (PP) especially in children and adolescents which ranges from 2.9–27%.^5^

It is said that the rate of polypharmacy is more prevalent among elderly patients and people with mental health problems, both of which are particularly at risk of adverse outcomes.^2^

Polypharmacy in psychiatry is an understudied area in Africa as few results are available for use. While antipsychotic polypharmacy practices are observed with growing frequency by prescribers, few studies have defined patient behaviors and treatment behaviors associated with the long-term use of this treatment strategy.^6^

This, therefore, necessitated the investigation of the prevalence and prescribers’ knowledge of Psychotropic Polypharmacy in the Bono and Ahafo Regions of Ghana.

## Materials and Methods

### Study Design

A quantitative study using descriptive parameters was employed to ascertain the prevalence and prescribers’ views and the effects of psychotropic polypharmacy in the Bono, Bono East, and Ahafo regions of Ghana. A descriptive study deals with the collection of data meant to explain or predict the conditions or relationships that may exist, opinions of subjects as well as practices that are going on. Data was collected between October and November 2020.

### Inclusion and exclusion criteria/indicators

Patients who were diagnosed with psychological disorders including psychotic, mood, and anxiety disorders were included in the study. Prescription charts (secondary data) across mental health units within these regions were reviewed retrospectively (data from patients’ recent or last three review visits were used) to identify all patients diagnosed with psychotic conditions to determine those who were co-prescribed with two or more antipsychotics. 250 patients’ folders (both inpatients and outpatients) within the respective mental health units were reviewed randomly but systematically (documented continuous prescription) from 1^st^ January 2019 through to 30^th^ March 2020. Two (2) to eleven (11) or more medications prescribed to a patient within the same time of interaction with the prescriber with the use of medication(s) from the same drug category or class.

Only qualified and permanent staff (prescribers) working within the region were randomly selected to respond to the questionnaire. Therefore, national service and rotational staff were excluded from this study. Also, mental health units that do not record a daily average OPD attendance with an average of 10 patients were not included in this study. Only subjects undergoing either pharmacotherapy alone or both pharmacotherapy and psychotherapy were used for this analysis. Psychotropic polypharmacy was regarded only if two or more psychotropic drugs (i.e., antipsychotics, anticholinergics, antidepressants, mood stabilizers, stimulants, etc.) are used during the same treatment time/concurrently (i.e., same class, multi-class, adjunctive and increase polypharmacy), not complete polypharmacy.

### Sample size and sampling method

The study utilized 309 patient folders and 58 prescribers.

To obtain the needed data for the study objectives, a random sampling method was used to select patients’ folders. However, a purposive sampling method was used to select the mental health units/facilities for data collection within the Bono, Bono East, and Ahafo regions of Ghana. Sample (patients’ folders) distribution and the selection were purposefully done based on size and Out Patient Department (OPD) attendance rate (average 10 patients daily) of the selected facilities as follows; Bono regional hospital: 80, Abrefi Hospital, Techiman 40, Kintampo government hospital 30, Goaso government hospital 40, Bechem government hospital 20, Berekum hospital 40 and Dorman Presbyterian hospital 40.

Prescribers were randomly selected proportionally at the respective Mental Health Units based on the number per facility as follows; Bono regional hospital: 15, Abrefi Hospital-Techiman: 5, Kintampo government hospital: 7, Berekum Hospital: 3, Wamfie government hospital: 4, Dormaa Presbyterian hospital: 4, Goaso government hospital: 7, Bechem government hospital: 3 and Sunyani municipal hospital:10.

The random sampling method was adopted because it takes a small proportion of the entire population to represent the entire data set ensuring that each subject has an equal chance of being selected for the study.

### Analysis of Data

Data collected from the field was analyzed with the aid of SPSS (version 25) using Stata version 6.0 for entry and analysis. Results are presented in tables, graphs, and charts representing the major findings relative to the objectives of this study.

### Ethical consideration

The Committee on Human Research and Publication Ethics (CHRPE), School of Medical Sciences, KNUST accordingly approved and gave ethical clearance as the protocol for the conduct of this study which bears the reference number CHRPE/AP/325/20.

The needed ethical clearance or approval was also sought from the respective hospital or unit heads before conducting this research via the Bono and Ahafo regional health directorates. The respondents were allowed to stay off from the exercise at any point in time. The participants were assured of anonymity, confidentiality, privacy, beneficence as well as non-maleficence. Thus, I assured participants that the exercise was purely academic and that it would be of importance to them and would not cause any of form harm to them. Hence, no participant was coaxed or forced into partaking in this study. They were also told not to provide any traceable personal information on the tool.

**Table 1:** Demographic characteristics of clients.

| Variable | Freq. (N=309) | Percentage (%) |
| --- | --- | --- |
| ***Age groups*** |  |  |
| 10yrs & Below | 11 | 3.6 |
| 11-20yrs | 52 | 16.8 |
| 21-30yrs | 84 | 27.2 |
| 31-40yrs | 77 | 24.9 |
| 41-50yrs | 40 | 12.9 |
| 51-60yrs | 33 | 10.7 |
| 61yrs & Above | 12 | 3.9 |
| **Mean age (STDV.)** | **33yrs (14.6)** |  |
| ***Gender*** |  |  |
| Male | 185 | 59.9 |
| Female | 124 | 40.1 |
| ***Facility*** |  |  |
| Berekum Health Center | 37 | 12.0 |
| Dormaa MHD | 28 | 9.1 |
| Goaso Government Hospital | 41 | 13.3 |
| Kintampo Municipal Hospital | 28 | 9.1 |
| Regional Hospital-Sunyani | 131 | 42.4 |
| Techiman polyclinic | 44 | 14.2 |

**Source: Fieldwork (2020)**

## Results

This study was conducted among two categories of respondents which were clients (data collected using their respective folders) and prescribers (mental health workers). Respectively, there were 309 clients and 58 prescribers across the study who willingly took part in this study. With 33 years as the mean age and a standard deviation of 14.6 among the clients, a lot of them were between the ages of 21-30 years. Only a few (3.6%) were below 10yrs. More than half of the respondents (clients) were males (59.9%) with the remaining as females.

**Table 2:** Demographic characteristics of the respondents (prescribers)

| **Variable** | **Freq. (N=58)** | **Percentage (%)** |
| --- | --- | --- |
| ***Age groups*** |  |  |
| 20 -30yrs | 35 | 60.3 |
| 31 - 40yrs | 22 | 37.9 |
| 41 - 50yrs | 1 | 1.7 |
| ***Gender*** |  |  |
| Male | 36 | 62.1 |
| Female | 22 | 37.9 |
| ***Marital Status*** |  |  |
| Married | 28 | 48.3 |
| Single | 30 | 51.7 |
| ***Qualification*** |  |  |
| RMN | 36 | 62.1 |
| CMHO | 21 | 36.2 |
| Others | 1 | 1.7 |

**Source: Fieldwork (2020)**

Among the prescribers, the majority were within the age group of 21-30 years. The majority of the respondents were males (62.1%) like the clients’ group. About half of them (51.7%) were singles whilst the remaining were married prescribers. More than half (62%) of them were registered mental health nurses (s) as per their qualifications.

### Prevalence of psychotropic polypharmacy

The study investigated clients’ folders (prescribing records) to find out how often PP occurs. This was done by identifying and categorizing clients based on the number of medications prescribed to them in each facility visited. These groupings were then used to ascertain the prevalence of PP as indicated in the chart below.

Findings from figure 1 above indicate that the majority (66%) of clients received 2 or more psychotropic drugs from prescribers at a time. On the other hand, in about 34% of data from prescription records, patients were given 1 drug after they were diagnosed.

**Figure 1:**
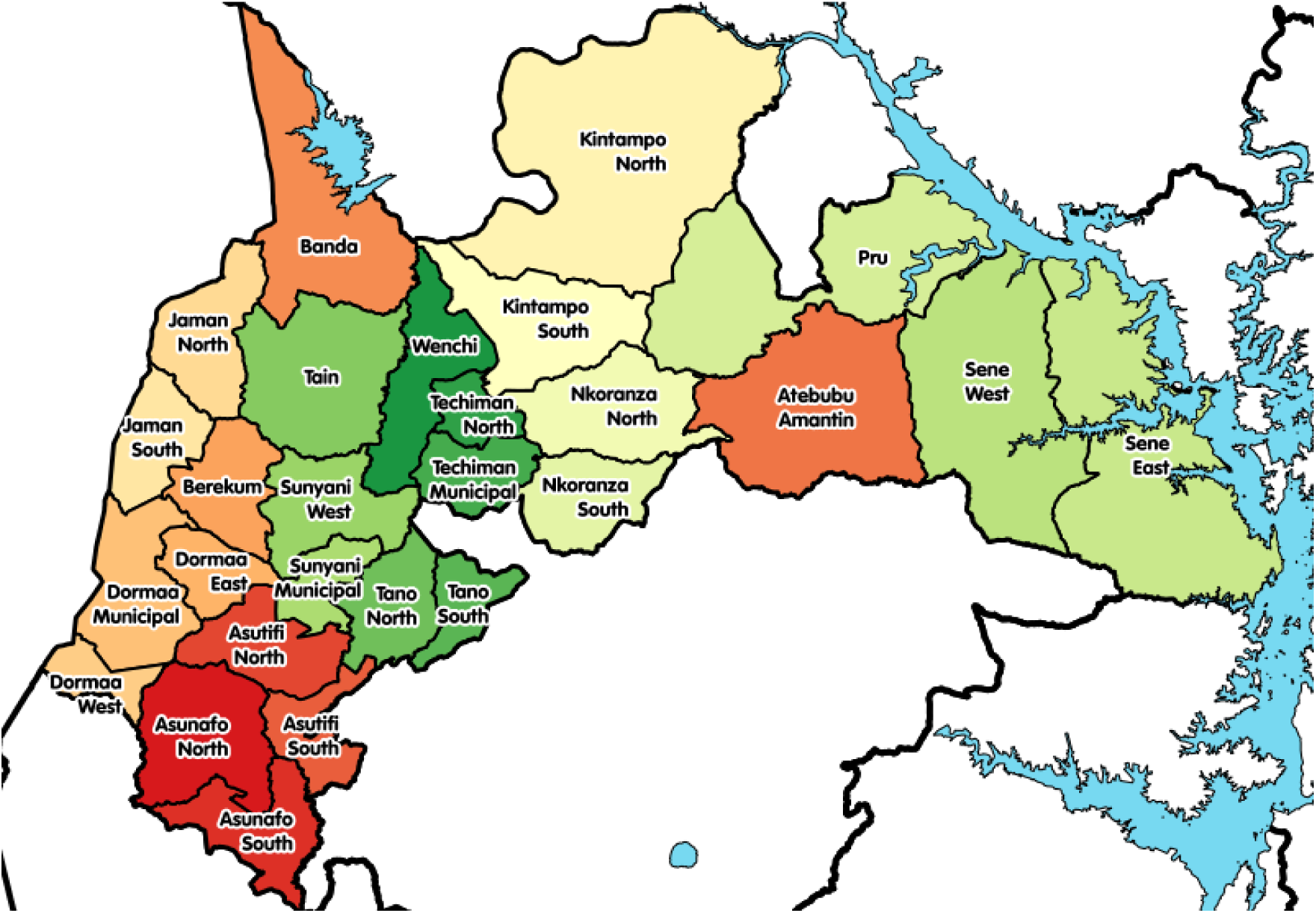
Map of Bono, Bono East, and Ahafo Regions of Ghana.

**Figure 2:**
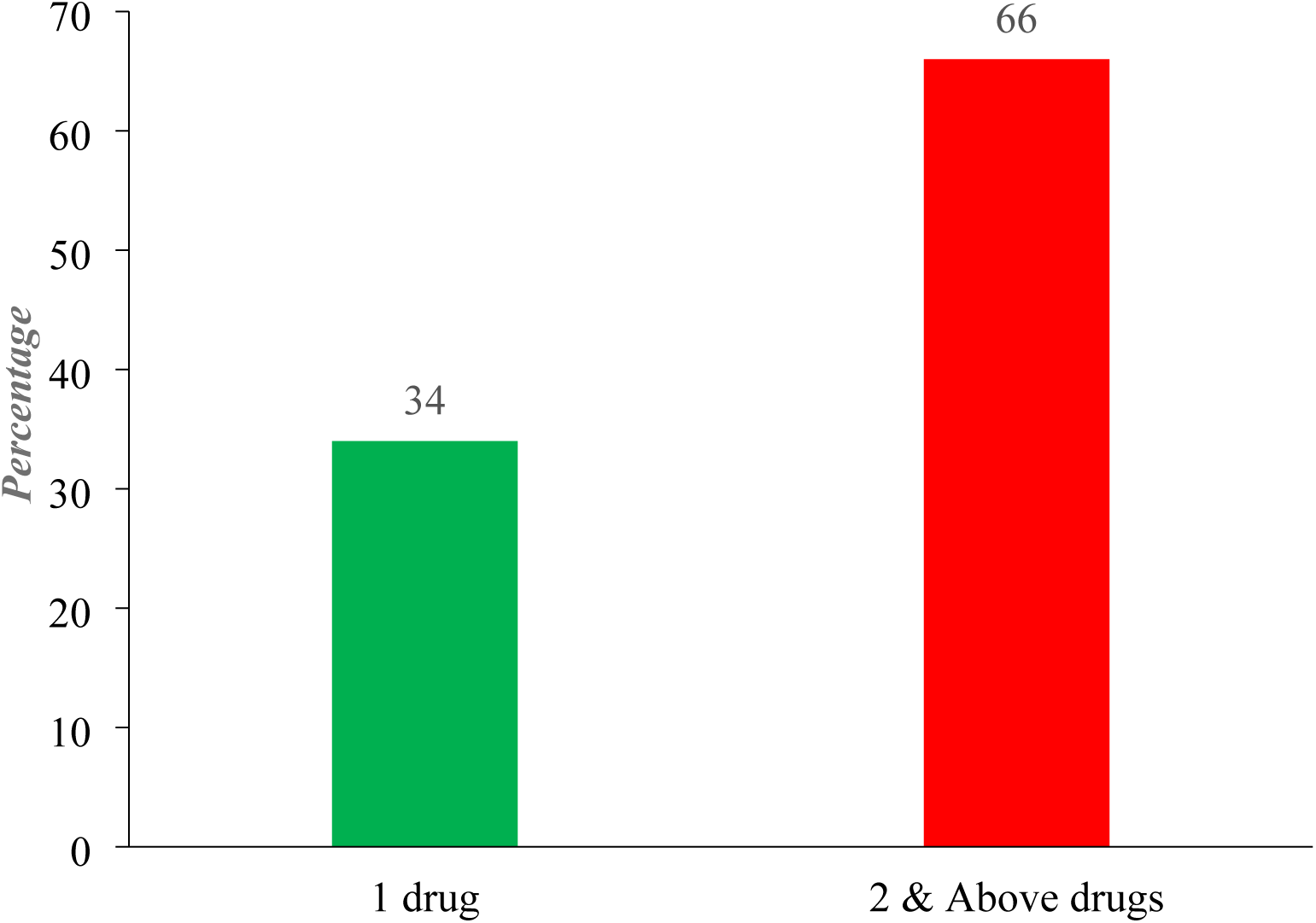
Prevalence of Psychotropic Polypharmacy (PP) Source: Fieldwork (2020).

Aside from the PP prevalence above, the study as well also revealed the number of medications given to clients after diagnosis in various categories as indicated in the table below;

**Table 3:** Quantity of drugs received by clients.

| **Number of drugs** | **Freq. (N=309)** | **Percentage (%)** |
| --- | --- | --- |
| 1 drug | 105 | 34.0 |
| 2 drugs | 72 | 23.3 |
| 3 drugs | 45 | 14.5 |
| 4 drugs | 24 | 7.8 |
| 5 drugs | 60 | 19.4 |
| 6 drugs & above | 3 | 1.0 |

**Source: Fieldwork (2020).**

Results from the table above indicate that about 105 clients out of 309 (representing 34%) received 1 drug after diagnosis. Clients who took 2 psychotropic drugs were as well 72 representing 23.3%. Also, clients who were prescribed 5 drugs were 19.4%. Clients who took 6 or more psychotropic medications were the last group of only 1%.

### Category of psychotropic medications frequently prescribed

The study seeks to investigate the types or specific categories of psychotropic medications that are commonly prescribed to clients. These psychotropic medications that were given to clients in this study were therefore classified into various categories which are indicated in the table below;

**Table 4:** Frequently prescribed psychotic medications.

| **Drug categories** | **Freq. (N=309)** | **Percentage (%)** |
| --- | --- | --- |
| Antipsychotics | 230 | 74.4 |
| Antidepressants | 29 | 9.4 |
| Mood stabilizers | 18 | 5.8 |
| Anti-anxiety | 1 | 0.3 |
| Anti-convulsant | 31 | 10.0 |

**Source: Fieldwork (2020).**

Results from the study indicate that the majority (74.4%) of the psychotropic medications that were frequently prescribed were mostly antipsychotic medications. Also, 10% of the prescribed medications were anti-convulsant medications. In contrast, the least prescribed medication from the study was identified to be anti-anxiety medications which were only 0.3% as shown above.

Moreover, the study also probed to ascertain which specific drug of the anti-psychotic class was frequently or mostly prescribed, findings on this are indicated in the figure 3 below.

**Figure 3:**
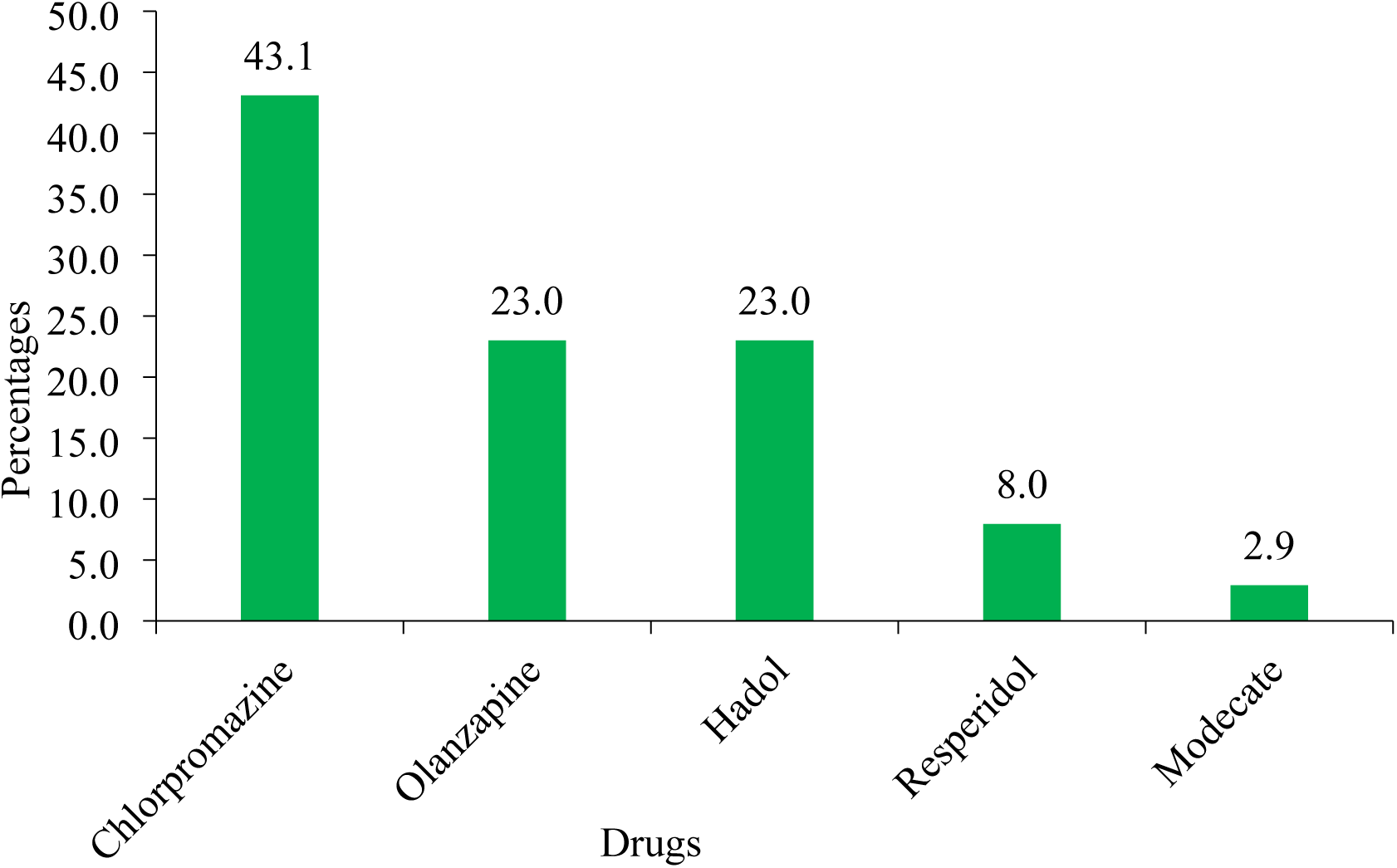
Antipsychotic prescription Source: Fieldwork (2020).

Results from the figure above show clearly that, close to half (43.1%) of the antipsychotic medications commonly issued were Chlorpromazine. Moreover, Olanzapine was the next most frequent anti-psychotic medication prescribed with 23%. As indicated in figure 3 above, injection Modecate (2.9%) was the least prescribed anti-psychotic medication among them.

### Concurrent combination of frequently prescribed antipsychotic medications

The study also investigated how often prescribers (concurrent/simultaneously) or co-prescribed these anti-psychotic medications as per client per visit. The results from this analysis are indicated in figure 4 below;

**Figure 4:**
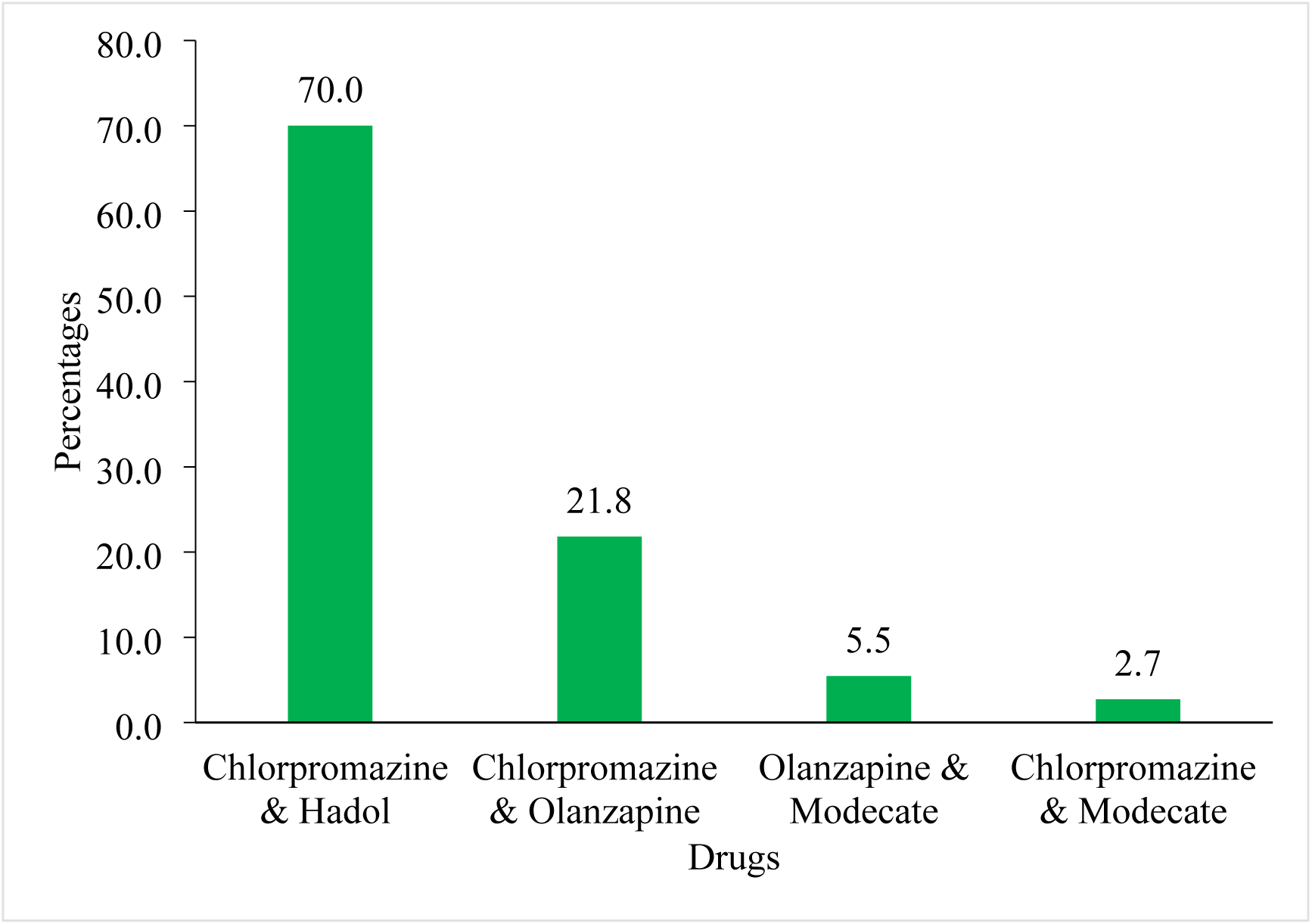
Combination of antipsychotic prescription Source: Fieldwork (2020).

Findings from this study revealed that, out of the frequently combined prescribed anti-psychotic medications, the majority (70%) combined Chlorpromazine & Hadol to clients. Also, about 21.8% of combined Chlorpromazine & Olanzapine to clients. However, the last combination was identified to be Chlorpromazine & Modecate (2.7%) as shown in the figure 4 above.

It was also noted that, in most (about 80%) of this combined prescription of antipsychotic medications, the tablet Artane was simultaneously added to the clients.

### Knowledge of prescribers (mental health workers) on psychotropic polypharmacy (PP)

The knowledge level (concerning PP meaning and its effects) of prescribers (mental health workers) was relevant to PP existence and therefore, this study examined their knowledge using the following variables in the table below.

**Table 5:**
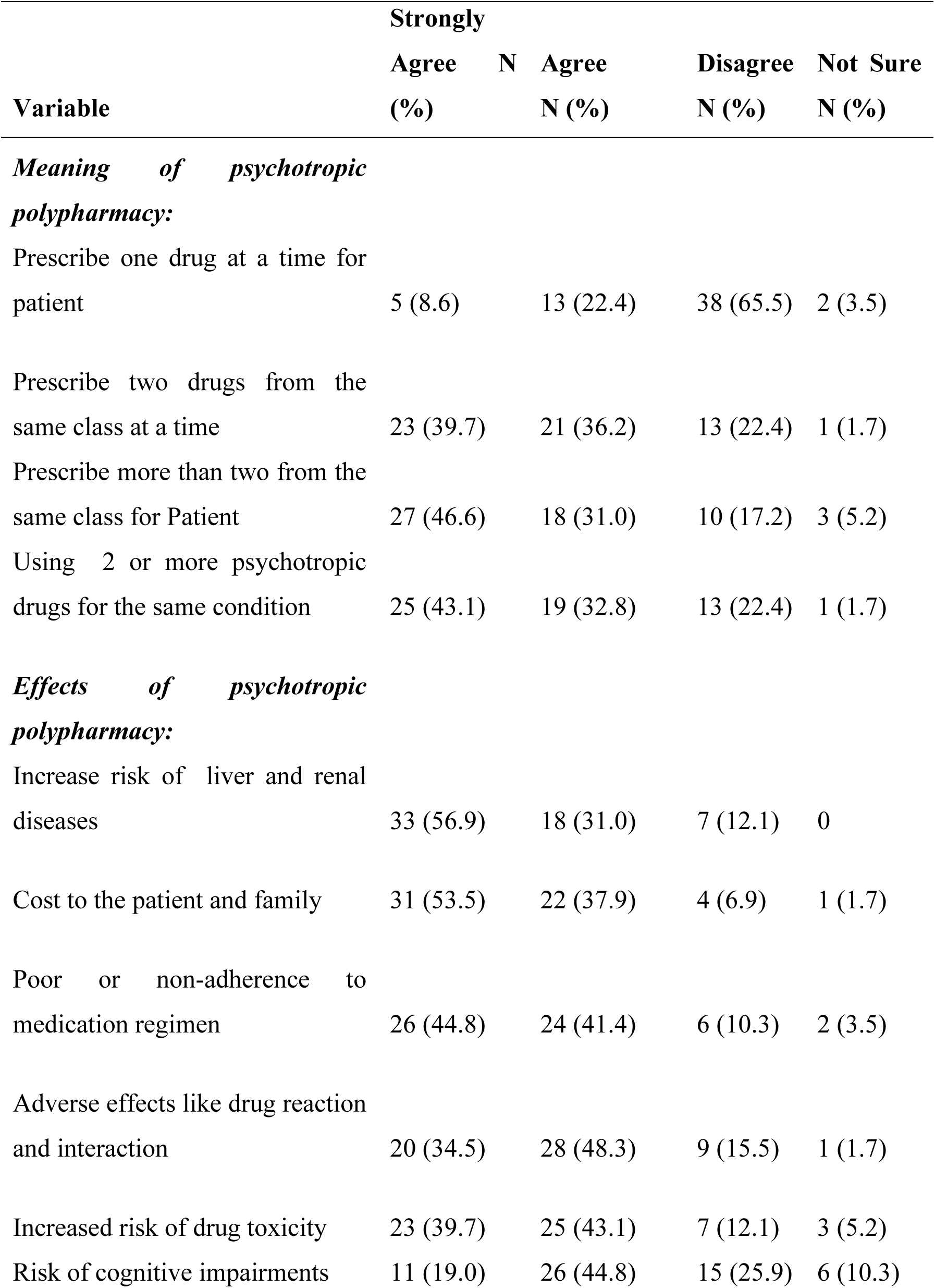
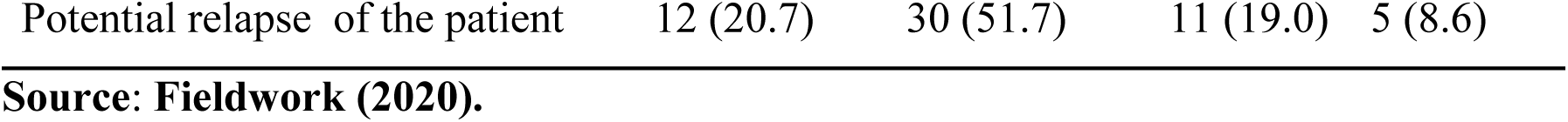
Knowledge of prescribers on psychotropic polypharmacy.

Results from the table above indicate that more than half (65.5%) of the respondents disagreed that prescribing only one drug to the client was PP. 22.4% of them agreed that it was polypharmacy prescribing one drug to clients whilst only 3.5% of the respondents were not able to tell if it was PP.

Again, respondents were asked if prescribing two drugs from the same category was considered PP. Results from the study indicated that more than one-quarter of the prescribers (39.7%) strongly agreed it was PP. On the contrary, 22.4% of them disagreed with this as being PP and only 1.7% of the prescribers were unable to tell if it was PP.

Prescribers were also asked if using two or more psychotropic drugs to treat the same condition equally constitutes PP. Findings from the study showed that close to half (43.1%) strongly agreed, 22.4%, disagreed with this, and 1.7% were not sure if this could be PP.

With prescribers’ knowledge relative to the effects of PP, the following were the respective responses to the statements made; on the extra cost to patients and families, about half (53.5%) strongly agreed. Contrarily, 6.9% of the respondents were also on the view that there is no cost, while 1.7% were unable to tell if that was an extra cost to patients and families.

Prescribers again were asked if polypharmacy could have a risk of cognitive impairment in patients. The results above showed that close to 44.8% agreed with this. On the contrary, 25.9% of the prescriber disagreed with any risk of cognitive impairment to the patient. About 10.3% were as well unable to tell if it could have this risk.

Moreover, 30% of the respondents agreed that PP can contribute to patient relapse whilst 11% disagreed.

### Rating of prescribers’ (mental health workers) knowledge of psychotropic polypharmacy

To determine their knowledge level, 11 questions that were measuring their knowledge based on responses were used to rate them. Respondents who scored 7 and above questions correctly were considered knowledgeable in PP. On the other hand, those who scored 6 and below correct were considered low knowledge of PP. This is indicated in the table below;

**Table 6:** Rating of prescribers’ knowledge of psychotropic polypharmacy.

| **Knowledge Rate** | **Frequency** | **Percentage (%)** |
| --- | --- | --- |
| *High (7-11)* | 48 | 82.8 |
| *Low (0-6)* | 10 | 17.2 |
| **Total** | **58** | **100.0** |

**Source: Fieldwork (2020).**

Findings from the study indicated that a greater majority (82.8%) of the prescribers scored between 7 and 11 correct, which indicated they had high knowledge of psychotropic polypharmacy. Contrarily, only 17.2% of them scored between 0 - 6 indicating low knowledge of psychotropic polypharmacy.

### Association between prescribers’ (mental health workers) knowledge level and their demographic characteristics

The study sought to ascertain if respondents’ (prescribers’) respective knowledge levels were influenced by their characteristics. A Pearson chi-square correlational test was carried out to find out this relation and the results are shown in the table below.

**Table 7:**
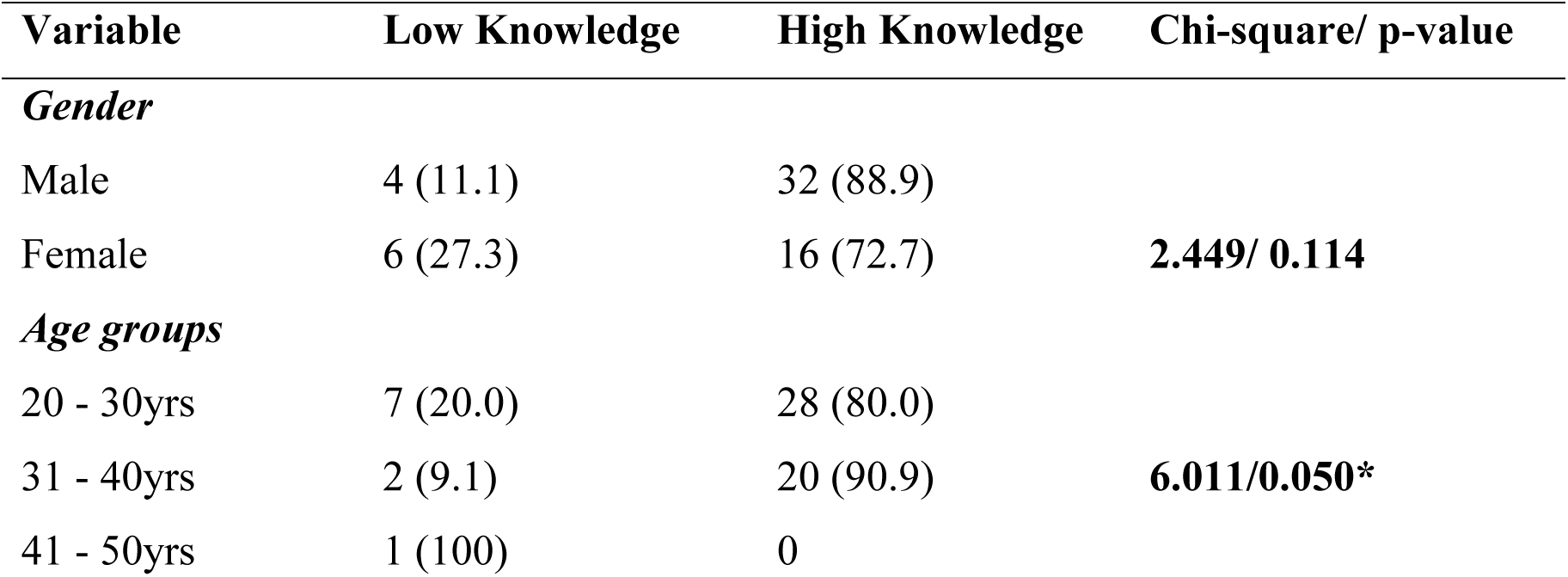
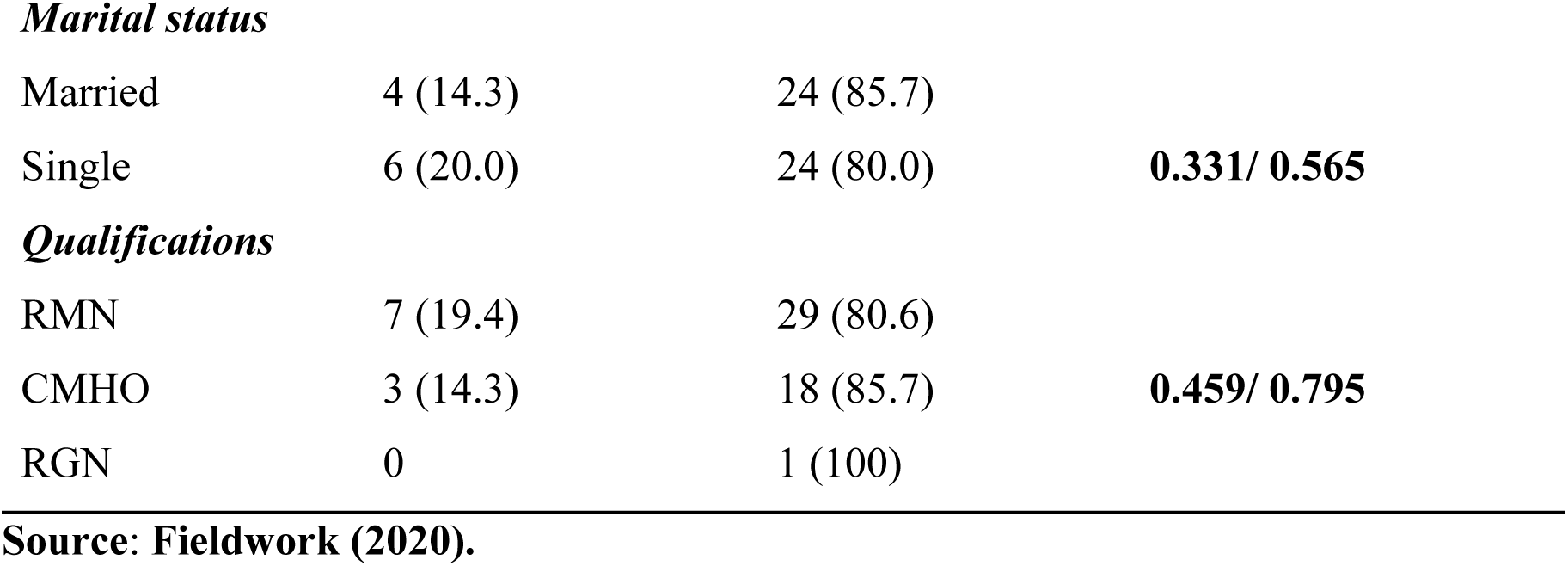
Association between prescribers’ knowledge level and demographic characteristics.

| Variable | Low Knowledge | High Knowledge | Chi-square/ p-value |
| --- | --- | --- | --- |
| *Gender* |  |  |  |
| Male | 4 (11.1) | 32 (88.9) | **2.449/ 0.114** |
| Female | 6 (27.3) | 16 (72.7) |  |
| *Age groups* |  |  |  |
| 20 - 30yrs | 7 (20.0) | 28 (80.0) | **6.011/0.050\*** |
| 31 - 40yrs | 2 (9.1) | 20 (90.9) |  |
| 41 - 50yrs | 1 (100) | 0 |  |
| ***Marital status*** |  |  |  |
| Married | 4 (14.3) | 24 (85.7) | **0.331/ 0.565** |
| Single | 6 (20.0) | 24 (80.0) |  |
| ***Qualifications*** |  |  |  |
| RMN | 7 (19.4) | 29 (80.6) | **0.459/ 0.795** |
| CMHO | 3 (14.3) | 18 (85.7) |  |
| RGN | 0 | 1 (100) |  |

**Source: Fieldwork (2020).**

Findings from the chi-square test indicated that respondents’ gender, marital status, and qualification were statistically not significant with their knowledge level as indicated in the table above. However, respondents’ ages were found to be statistically significant with their knowledge level (n=58, ꭓ2= 6.011, p=0.050, α=5%).

### Reasons prescribers (mental health workers) may consider PP as an option

This study as well seeks to investigate reasons why prescribers will consider PP as another option in treating their patients/clients. A series of questions were used to probe this as indicated in the table below.

**Table 8:**
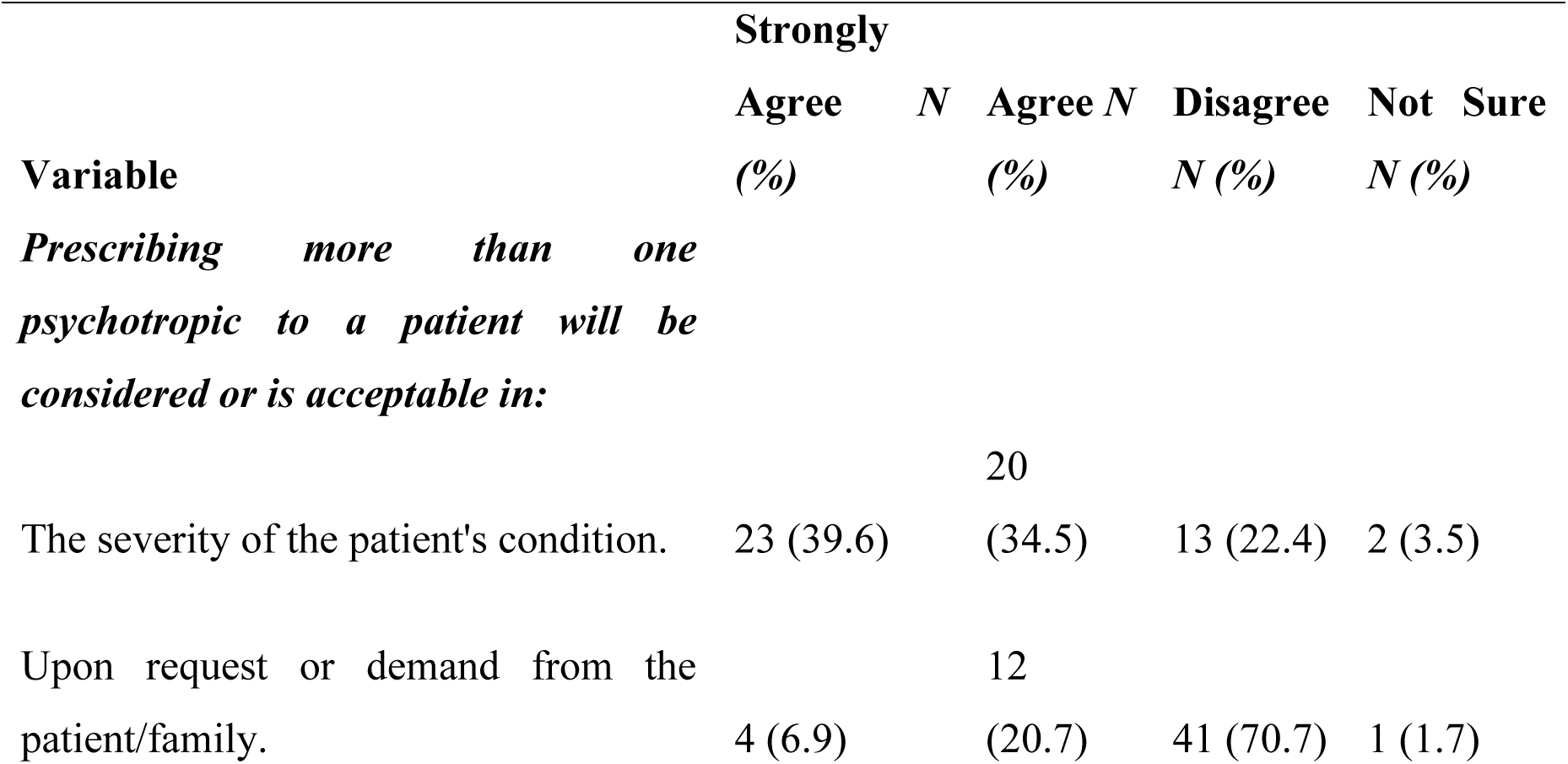
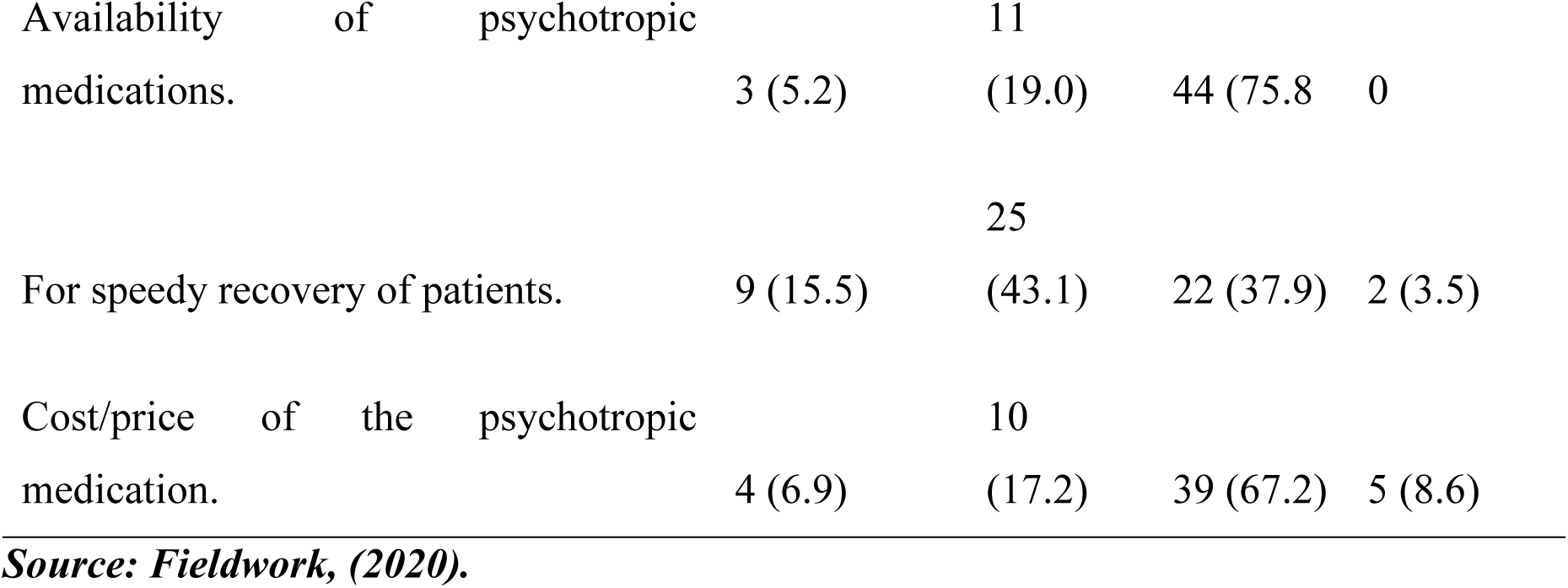
Reasons for considering psychotropic polypharmacy.

Findings from the study revealed that 39.6% of prescribers strongly agreed that PP should be an option when the patient’s condition is severe. On the contrary to this 22.4% also disagreed with that fact, the severity of the condition should not lead to polypharmacy. However, 3.5% were unable to tell if that could lead them to take that decision.

Again, respondents were asked if drug availability could usually influence their practice of PP at any time. The results indicate that the majority (75.8%) disagreed with this as a pre-condition for PP practice. Only 19% agreed that PP should be practiced under this circumstance.

Respondents were also asked if the speedy recovery of patients (expected desire) would influence the practice of PP. The results indicated that close to half (43.1%) of the respondents confirmed that could happen. However, 37.9% disagreed with the fact that speedy recovery does not or should not warrant PP and 3.5% were not sure if this could be a factor.

### Prescribers’ view on the encouragement or promotion of psychotropic polypharmacy in psychiatry

**Table 9:**
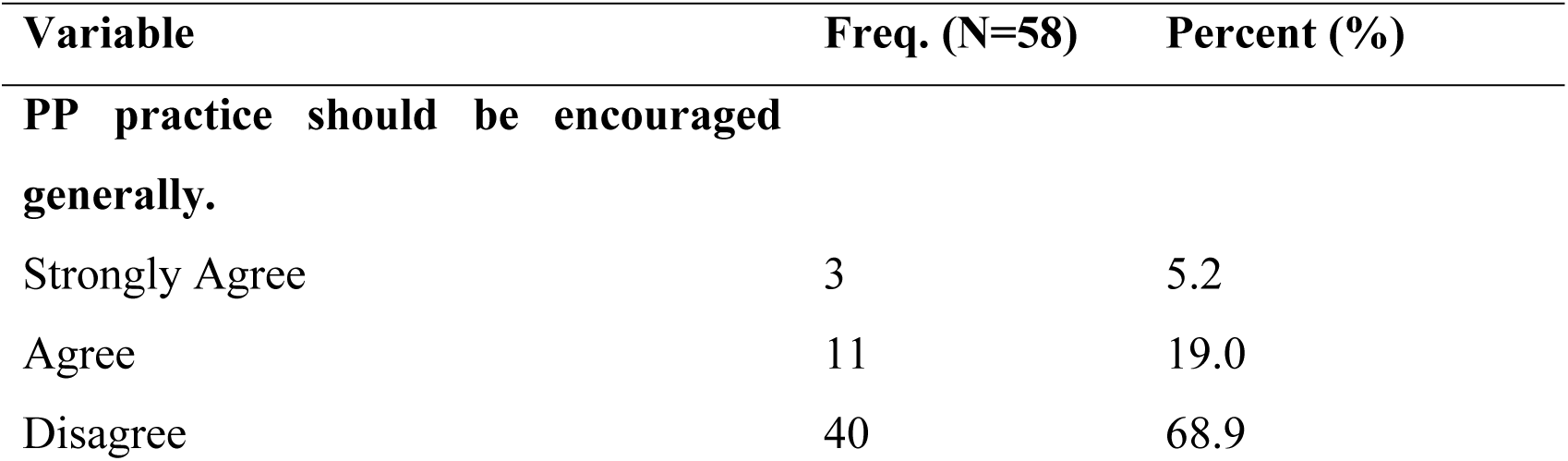
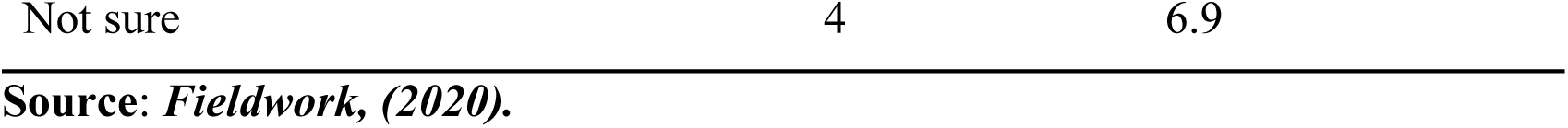
Views of prescribers.

With the idea of encouraging psychotropic polypharmacy, more than half (68.9%) of the respondents disagreed that PP should be encouraged or promoted in the treatment of patients. However, 19% of the respondents supported that idea or statement whilst 6.9% were unable to tell if it should be encouraged or not.

### Other findings from the study

The study investigated respondents’ diagnoses concerning frequently prescribed psychotropic polypharmacy and came out with the following results as indicated in the figure below.

**Figure 5:**
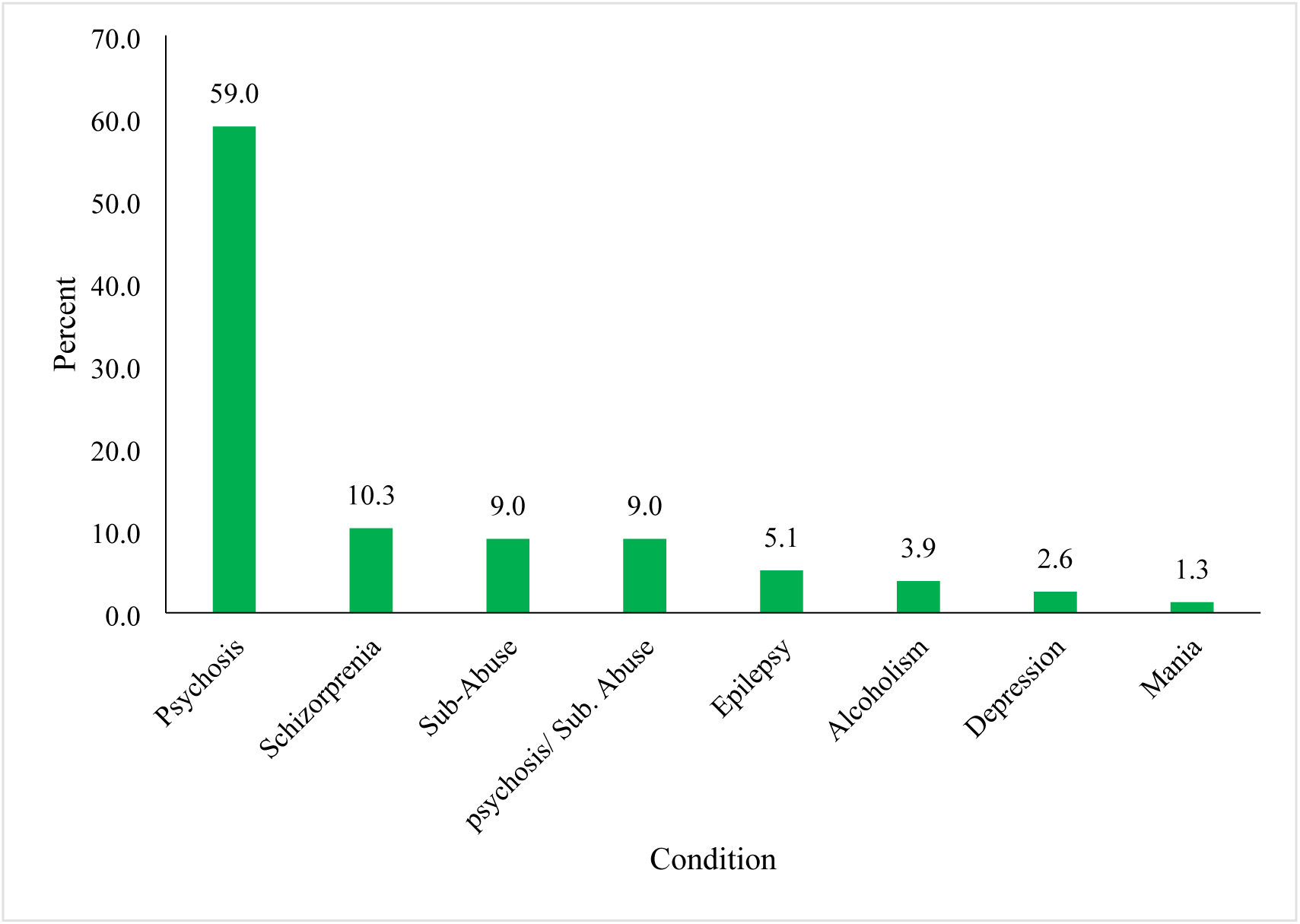
prescription pattern relative to condition Source: Fieldwork (2020)

Results from the study show that the frequently occurring polypharmacy which is Chropromazine & Hadol was prescribed to the majority of patients suffering from psychosis, followed by patients diagnosed with schizophrenia. On the other hand, only 1.3% of patients diagnosed with mania were involved in PP.

### Age and Psychotropic Polypharmacy

The study also found out the age group of patients that was more involved in PP and the findings are shown in the table below;

**Table 10:**
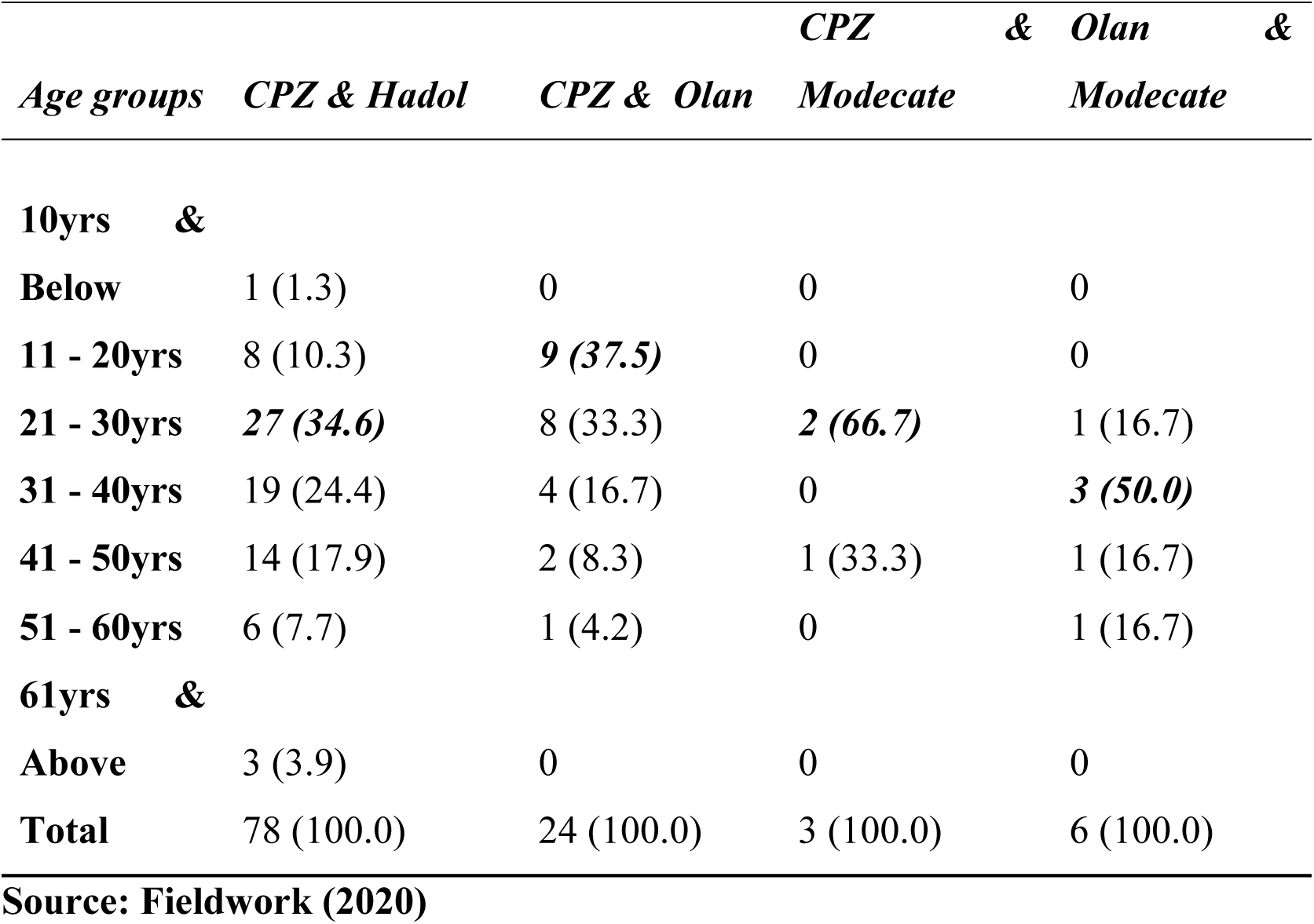
Age and Psychotropic Polypharmacy.

| *Age groups* | *CPZ & Hadol* | *CPZ & Olan* | *CPZ & Modecate* | *Olan & Modecate* |
| --- | --- | --- | --- | --- |
| **10yrs & Below** | 1 (1.3) | 0 | 0 | 0 |
| **11 - 20yrs** | 8 (10.3) | **9 (37.5)** | 0 | 0 |
| **21 - 30yrs** | **27 (34.6)** | 8 (33.3) | **2 (66.7)** | 1 (16.7) |
| **31 - 40yrs** | 19 (24.4) | 4 (16.7) | 0 | **3 (50.0)** |
| **41 - 50yrs** | 14 (17.9) | 2 (8.3) | 1 (33.3) | 1 (16.7) |
| **51 - 60yrs** | 6 (7.7) | 1 (4.2) | 0 | 1 (16.7) |
| **61yrs & Above** | 3 (3.9) | 0 | 0 | 0 |
| **Total** | 78 (100.0) | 24 (100.0) | 3 (100.0) | 6 (100.0) |

**Source: Fieldwork (2020)**

Results from the table above show that more than one quarter (34.6%) of the patients aged between 21-30 years were given the combination of Chropromazine & Hadol with about 1.3% of patients who were 10 years and below taking this same combination. Again, the “CPZ & Olanzapine” combination was given to patients between the ages of 11-20years. The results also show that “CPZ & Modecate” combined prescription to patients occurred mostly among respondents aged 21-30 years (66.7%). About 50% of the “Olanzapine & Modecate” prescription was among the 31-40 years of age group.

### Sex and Psychotropic Polypharmacy

Furthermore, additional probing was done to find out which sex group of patients received these categories of polypharmacy and the table below shows the findings.

**Table 11:**
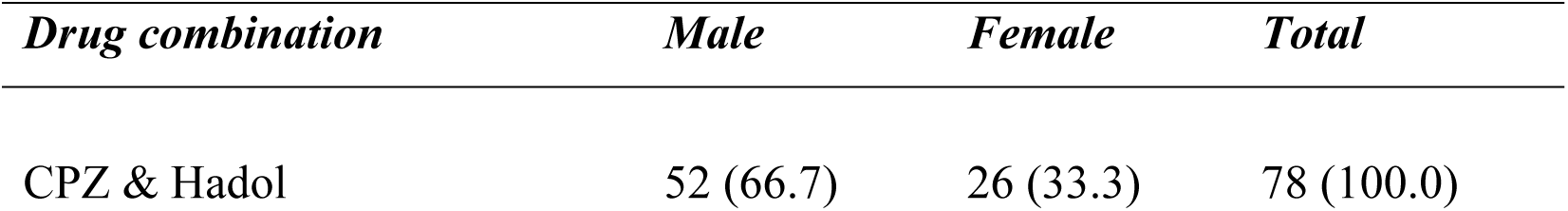
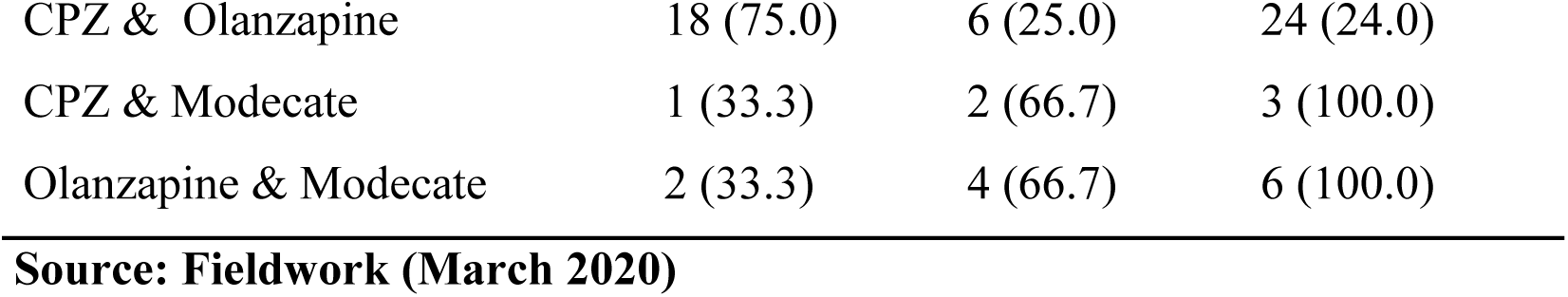
Sex and psychotropic polypharmacy.

| *Drug combination* | *Male* | *Female* | *Total* |
| --- | --- | --- | --- |
| CPZ & Hadol | 52 (66.7) | 26 (33.3) | 78 (100.0) |
| CPZ & Olanzapine | 18 (75.0) | 6 (25.0) | 24 (24.0) |
| CPZ & Modecate | 1 (33.3) | 2 (66.7) | 3 (100.0) |
| Olanzapine & Modecate | 2 (33.3) | 4 (66.7) | 6 (100.0) |

**Source: Fieldwork (March 2020)**

Results from the table above showed that more than half (66.7%) of the CPZ & Hadol combined prescription was among male patients with only 33.3% among females. Also, the majority of the “CPA & Olanzapine” polypharmacy occurred within male patients. However, more than half (66.7%) of both “CPZ & Modecate” and “Olanzapine & Modecate” polypharmacy occurred among female patients

## Discussion

### Prevalence of Psychotropic Polypharmacy

A study in Nigeria reported a high prevalence of psychotropic polypharmacy (PP) ranging from 2.9 – 27%, especially in children and adolescents.^5^ In this study, the prevalence of psychotropic polypharmacy was assessed by identifying and categorizing clients according to the number of medications prescribed to them in each facility under study. The study found that only 34% of clients who visited the facility were prescribed one drug while 66% of clients were prescribed two or more psychotropic drugs. The breakdown of the 66% of clients prescribed 2 or more drugs revealed that 23% were prescribed exactly 2 drugs while 19% were prescribed 5 drugs, 14% were prescribed 3 drugs with 7.8% and, 1% prescribed 4 and 6 drugs respectively. This indicates that clients have a high probability of receiving more than two drugs on a visit to the facility.

The finding of this study corroborates with another study which found that the prevalence of polypharmacy in psychiatry varies between 13%–90%.^7^ Similar studies in Europe also found that the prevalence of polypharmacy ranges between 33.3% to 73.3. % depending on the adopted definition.^8^ Another study of a cohort of elderly people in Bambuν city to evaluate the prevalence of polypharmacy and the influence of income on the association between medication use and cognitive impairment among elderly people.^9^ They reported that within the geriatric age group (age > 60 years), 44.8% of the 1,554 elderly Bambuν cohorts were consuming 2-4 medications and 25.5% were consuming five or more medications.

The findings of this study also revealed that patients aged between 21-30 years were more (34.6%) involved in PP and received CPZ+ Hadol and CPZ+ Modecate combinations, but the combined prescription of Olanzepine+ Modecate was high (50%) among patients aged between 31-40years.On the other hand patients, 61 years who were more likely to be diagnosed with dementia received less PP. This confirms the findings of another study that younger and unmarried patients were more likely to receive PP, hence exposing them to all the associated risks of PP.^11^

### Category of psychotropic medications frequently prescribed in PP

A study using a nationwide representative sample of 3,466 children and adolescents, reported the prevalence of multi-class psychotropic treatment to be 19% in this population. Antidepressants were the most common co-prescribed medication class in multi-class visits followed by ADHD medications, antipsychotics, mood stabilizers, and sedative-hypnotics. ^12^

The study employed five categories to group the types of psychotropic drugs frequently prescribed; anti-psychotics, mood stabilizers, anti-depressants, anti-anxiety, and, anti-convulsants. Anti-psychotic drugs were found as the most (74.4%) prescribed category of medication, this corroborates the findings by another study which found that antipsychotic polypharmacy is widely prevalent and all available evidence suggests that antipsychotic polypharmacy is common in clinical practice.^13^

Anti-convulsant were 10% of the prescribed category, anti-depressants were 9.4%, with 5.8% and 0.3% for mood stabilizers and anti-anxiety respectively. These percentages indicate that clients are the least prescribed drugs from these categories in the facilities under this study. This finding is contrary to an earlier study which found that 6 % of patients received typical, while 39 % received atypical antipsychotics.^14^ There was greater use of antidepressants (23 %), mood stabilizers (19 %), and antianxiety agents (16 %) relative to other studies. The use of anti-impulsive drugs, stimulants, and, hypnotics was rare (1–2 %).

A study found that half of the patients used in the study received combinations of non-clozapine oral antipsychotics and the other half were receiving either clozapine or an injectable antipsychotic.^15^ The findings of this study found that the concurrent combination of frequently prescribed anti-psychotic medications indicated that, the prescription of Chlorpromazine with Hadol was highest 70%, Chlorpromazine with Olanzapine (21.8%), Olanzapine with Modecate (5.5%), and Chlorpromazine with Modecate 2.7%. This finding in this study could support the general idea that most patients potentially experienced side effects of their medications including EPSE as the two most combined prescribed antipsychotic drugs are known and reported to have such potentials comparatively.

### Knowledge of prescribers (mental health workers) in PP

Polymedicine or polytherapy is explained as the use of multiple medications prescribed appropriately for treating multiple comorbid conditions.^16^ Polypharmacy is a simple term that can be said to be the prescription and use of many drugs at a time to manage a condition. The argument about polypharmacy is not solely about the number of medications used, but also about the effectiveness, utility, and potential harm of each medication, either individually or in combination.^3^ The knowledge of the prescriber and pharmacy workers is very important because it influences the prescriptions given (pattern) by a prescriber.^17^

The study sampled 58 prescribers, 36 (62.1%) are RMN, 21(36.2%) CMHO, and 1(1.7%) other health practitioners. In the assessment of the knowledge of prescribers on the meaning of polypharmacy, 65.5% of prescribers disagreed that prescribing only one drug to the client was polypharmacy. 39.7% of prescribers agreed the prescription of two drugs from the same category as polypharmacy, 22.4% disagreed and 1.7% were unable to tell. 43.1% of prescribers indicated using 2 or more psychotropic medications to treat the same condition equally constitutes PP, with 22.4% not in agreement and 1.7% not sure if this could be PP. Prescribers° knowledge relative to the effects of PP, about half (53.5%) strongly agreed on the extra cost to patients and families. Contrarily, 6.9% of the respondents were also on the view that there is no cost, while 1.7% were unable to tell if that was an extra cost to patients and families.

When prescribers were asked if polypharmacy could have a risk of cognitive impairment in patients, close to 44.8% agreed to this. On the contrary, 25.9% of the prescriber disagreed with any risk of cognitive impairment to the patient. About 10.3% were as well unable to tell if it could have this risk. Moreover, 30% of the prescribers agreed that PP can contribute to patient relapse whilst 11% disagreed.

A systematic study revealed that there were 111 numerical-only definitions of what polypharmacy means.^16^ In all 80.4% of the definitions, 15 quantity definitions that incorporated a duration of therapy or healthcare setting represented 10.9% and 12 descriptive definitions as 8.7%. The study also revealed that the most commonly reported definition of polypharmacy was the numerical definition of five or more medications daily (n = 51, 46.4% of articles), with definitions ranging from two or more to 11 or more medicines. Only 6.4% of articles were observed to have classified the variation between appropriate and inappropriate polypharmacy, which is using descriptive definitions to make this distinction.

### Rating of prescribers’ (mental health workers) knowledge of PP

The lack of a consistent, clear, and globally accepted definition of polypharmacy including terms such as minor, moderate, and major polypharmacy, has made it pretty difficult for healthcare professionals to assess and consider efficacy and safety issues within the clinical setting.^18^ A study indicated that most nurses do have the ability to assess the side effects of psychotropic medications, hence are capable of doing so relative to psychotropics poly-pharmacy.^17^

Prescribers were rated to ascertain their overall knowledge of psychotropic polypharmacy based on their score in answering the 11 questions which were measured knowledge-based on responses. Respondents who scored 7 and above questions correctly were considered knowledgeable in PP and those who scored 6 and below correct were considered low knowledge on PP. It was found that 82% were highly knowledgeable and 17.2% had low knowledge in polypharmacy.

These findings however did not support the findings of the study which found that hospital pharmacists had low knowledge of psychotropic polypharmacy.^17^

### Association between prescribers’ (mental health workers) knowledge level and their demographic characteristics

The study investigated the association between prescribers’ (mental health workers) knowledge level and their demographic characteristics. Findings from the chi-square test indicate that respondents’ gender, marital status, and qualification were statistically not significant with their knowledge level. However, respondents’ ages were found to be statistically significant with their knowledge level (n=58, ꭓ2= 6.011, p=0.050, α=5%) which can be linked to their level of experiences gathered over the years on the field.

### Reasons prescribers (mental health workers) may consider PP as an option

A study indicated that psychotropic polypharmacy is still a common practice despite inconclusive scientific evidence of additional efficacy and potential exacerbation of undesirable effects of medications.^19^ A study on some reasons for polypharmacy practice stated antipsychotic polypharmacy (APP) might be appropriate in patients who have failed or cannot tolerate antipsychotic monotherapy put forward.^12^ The safety data for psychotropic polypharmacy are mixed; several studies of clozapine plus risperidone or adjunctive aripiprazole suggest that these combinations generally are well tolerated with positive effects among patients

Findings from this study revealed that 39.6% of prescribers strongly agreed that PP should be an option when the patient’s condition is severe whereas 22.4% also disagreed severity of the condition should not lead to polypharmacy. However, 3.5% were unable to tell if the severity of the condition could lead them to take that decision. This means that prescribers engaged in PP because they anticipated a particular effect(s), especially to the patient who reported to their units.

## Other findings from the study

### Category of diagnosed patients and psychotropic polypharmacy

This study found out that, more patients (59%) who were diagnosed with psychosis were involved in PP as compared to patients with Schizophrenia/Schizo-affective disorder of 10.3%. These findings vary from that of an earlier study which indicated that patients diagnosed with Schizophrenia/Schizo-affective disorders were more likely to receive oral antipsychotics PP.^20^

### Age and psychotropic polypharmacy

The study revealed that patients who were between the age group of 21-30 years received more (34.6%) PP as compared to other age groupings. They received more combinations of CPZ+Hadol and 667% of CPZ+Modecate prescriptions. The co-prescription of Olanzapine + Modecate was common among the 31-40years age group. However, patients above 61 years received less PP. This finding is consistent with the findings of a previous study which indicated that younger and unmarried patients suffering from psychiatric conditions were more likely to receive PP comparatively, therefore, exposing them to all the potential negative effects of PP like poor and decline/lower cognitive functioning including EPSE of the drugs.^21^

### Sex and psychotropic polypharmacy

In this study, it was revealed that male patients who visited the respective facilities were more (66.7%) involved in PP with the combined prescription of CPZ+ Hadol than females. However, in PP instances of combined prescription of CPZ+Modecate, female patients were more dominant comparatively. These findings mean that male patients were at higher risk of experiencing EPSEs of drugs than female patients, for these combinations of drugs in PP is known to be among the high-risk drugs combination in PP as indicated by findings in a previous study.^22^

## Conclusions

The following major conclusions were drawn based on the main results of the study.

1. There is a high prevalence of PP in the study area and 2 or more psychotropic drugs were prescribed to a patient per visit (i.e. numerical PP). More patients diagnosed with psychosis were involved in PP and patients suffering from Mania were less involved in PP.
2. Prescribers within the study area had a high understanding of what PP was.
3. Same-class polypharmacy involving antipsychotic polypharmacy was the more common in the study area whilst anti-anxiety medications were least prescribed.
4. The combination of Chlorpromazine and Hadol (typical) was prevalent in the PP practice relative to antipsychotic polypharmacy and more males between the age group of 21-30 years received this combination than female patients.
5. The majority of the prescribers believed that PP in psychiatry should be discouraged but indicated that PP should be considered for the speedy recovery of the patient.

## Authors’ contribution

**AJB** and JD conceptualized the overall study with its goals and aims. MMI retrieved the requisite data from databases. SSA and LS played a supportive role in data consolidation. WA wrote the study background and played a supportive role in data analysis. FA played a role in designing the methodology. SA played a vital role in designing the methodology for the study. BPN played a role in writing the discussion of the study. JD supported the writing, review, and editing of the manuscript. AJB, MMI, and SSA did the data analysis for the study.

All authors contributed significantly to the critical revision and approved the final version before the onward submission.

## Data Availability

All relevant data are within the manuscript and its Supporting Information files.

## Acknowledgment

Authors express their profound gratitude to prescribers (mental health professionals) within the Bono and Ahafo regions who devoted their time and efforts to making the study a success.

